# Polymorphism of *NAT2, PXR, ABCB1*, and *GSTT1* genes among tuberculosis patients of North Eastern States of India

**DOI:** 10.1101/2021.06.09.21258600

**Authors:** Heikrujam Nilkanta Meitei, Anupama Pandey, Hossain Md. Faruquee, Maria Thokchom, Sonia Athokpam, Hritusree Guha, Ranjit Das, Sourav Saha, Rukuwe-u Kupa, Wetetsho Kapfo, Joshua Keppen, Amit Kumar Mohapatara, Haripriya Priyadarsini, Arunkumar Singh Koijam, Arunabha Dasgupta, Bidhan Goswami, Aseno Thong, Kezhasino Meru, Wungyong Konyak, Dinesh Gupta, Anjan Das, Vinotsole Khamo, Lokhendro Singh Huidrom, Sunita Haobam, Ranjan Kumar Nanda, Reena Haobam

## Abstract

**Background:** Anti-tuberculosis drug-induced liver injury (AT-DILI) in tuberculosis (TB) patients has been linked to polymorphisms in genes encoding drug metabolism enzymes and proteins.

**Objective:** This study aimed to monitor polymorphisms of *NAT2, PXR, ABCB1*, and *GSTT1* genes in TB patients from three states (Manipur, Tripura, and Nagaland) in the North Eastern Region of India.

**Methods:** Genomic DNA was isolated from the whole blood samples of TB patients (n=219; Manipur:139; Tripura: 60; Nagaland: 20). The TaqMan allelic discrimination assay and statistical tools were used to investigate single nucleotide polymorphisms (SNP) patterns in *NAT2, PXR, ABCB1*, and *GSTT1* genes.

**Results:** In the study population, ten distinct genotypes of the *NAT2* gene and single variation in the *PXR, ABCB1*, and *GSTT1* genes were identified. A strong linkage disequilibrium (LD) was observed between rs1801280 and rs1799931 of the *NAT2* gene. Majority of the study populations were intermediate (~46.1%), rest were either slow acetylators (~35.6%) or fast acetylators. Interestingly, ~55% of the TB patients in Tripura were slow acetylators and majority in Manipur and Nagaland were of intermediate acetylator genotypes. For all of the markers investigated, the population had a greater prevalence of ancestral alleles and genotypes. According to a combinational study of the genotypes linked to AT-DILI, ~26.1% of the population possessed the risk genotypes.

**Conclusion:** These TB patients from north eastern states of India were found as carriers of the ancestral alleles and genotypes. And the risk for AT-DILI during TB treatment is low. Expanding such studies with additional markers and larger sample sizes will be useful to generate precise population-specific pharmacogenomics details for efficient TB management.

## Introduction

Tuberculosis (TB) disease is caused by infection of *Mycobacterium tuberculosis (Mtb)* and leads to ~2 million annual deaths worldwide [1]. In India, the TB incidence rate is ~159/1,00,000 with a ~4% fatality rate [2]. Treatment strategy in TB patients involves multiple drugs for long duration up to 6-18 months depending on the drug susceptibility or resistance status of Mtb. Majority of these anti-TB drugs undergo further conversion by the host enzymes to become active molecules for effective killing of Mtb. This contributes to certain side effects like skin reactions, hepatitis, nausea, vomiting, purpura, lethargy, dizziness, abdominal pain, gastrointestinal and neurological disorders [3,4]. About ~1-47 % of the TB patients, during treatment, develop anti-tuberculosis drug-induced liver injury (AT-DILI) [1]. The type of prescribed drugs, race, age, genetic factors, sex, alcohol intake, TB condition, and co-infection with HIV, hepatitis B and C virus [5,6,7,8,9,10,11,12,13] influences the severity of AT-DILI. With discontinuation of the treatment, AT-DILI condition is also reversed [14]. Certain variations in the genomic regions within or near the genes encoding drug-metabolizing enzymes (*NAT2, CYP2E1, GSTs*), lipid metabolism (*CYP7A1, BSEP, UGTs, PXR*), immune adaptations (*HLAs* and *TNF-*α), transporter proteins (*ABCB1*) and oxidant challenges (*TXNRD1, SOD1, BACH1*) were reported to be associated with AT-DILI [15].

NAT2 is a cytosolic phase II conjugation enzyme responsible for the deactivation of isoniazid to acetyl isoniazid [16,17,18,19]. It further hydrolyses acetyl hydrazine, which is subsequently oxidized by cytochrome P450 2E1 (CYP2E1) to intermediates with hepatotoxic effect. *NAT2* gene, present on chromosome 8, is reported to show high polymorphisms with 36 allelic variants representing varied acetylation activities (rapid acetylators: RAs, homozygous of *NAT2*4* alleles; intermediate acetylators: IAs, heterozygous of *NAT2*4* alleles; and slow acetylators: SAs, homozygous/non-heterozygous of *NAT2*4* alleles) [20,21,22]. Most variants (single nucleotide polymorphisms: SNPs) of the *NAT2* gene are located within the 873bp intronless coding region. Four SNPs rs1801280 (341 T>C; *NAT2*5*), rs1799930 (590 G>A; *NAT2*6*), rs1799931 (857 G>A; *NAT2*7*), rs1801279 (191 G>A; *NAT2*14*) are associated with slow acetylator phenotype [23]. TB patients with low NAT2 enzyme activity show higher circulatory hydrazine levels creating a significant risk to develop AT-DILI [24, 25, 26, 27].

Similarly, *PXR* in the liver, regulates the expression of many genes (CYP3A4 isoenzyme, UGT1A1, MDR1, ABCB1, glutathione S-transferase (GST), SULT2A1, UGT1A1, MRP2, OATP2) involved in hepatic drug-clearance system [28,29]. Polar groups like glucuronides are added to anti-TB drugs for detoxification by these enzymes before elimination from the host [30]. Polymorphism in the *PXR* gene (rs3814055; C>T) has shown an association with an increased risk of AT-DILI [31].

Glutathione S-transferases (GSTs) are phase II drug-metabolizing enzymes involved in protecting cells from oxidative stress. Xenobiotics including drugs (like isoniazid and rifampicin) and reactive oxygen species (ROS) cause liver injury and GSTs conjugate glutathione to these groups before elimination to protect the liver from damage. *GSTM1* and *GSTT1*, members of *G*ST, are involved in the conjugation of isoniazid. Homozygous null mutation at these loci is reported to show a higher rate of AT-DILI [32, 33].

*ABCB1 (MDR1)* gene, located on human chromosome 7q21.12, is reported to be highly polymorphic [34, 35]. The SNP present in exon 26 (C3435T SNP: rs1045642), alters the expression of P-gp and it affects the drug pharmacokinetics of its substrate drugs [36, 37]. Subjects with the homozygous 3435 TT genotype of *ABCB1* is reported to show a higher (>3-fold) chance of developing DILI [38]. The population level genetic polymorphisms pattern varies significantly within and across countries. India harbors highest TB cases globally and presents a very diverse population with varied ethnicities. Importantly, very limited pharmacogenomics reports are available from the TB patients of the north eastern states of India that shares international boundary with Myanmar and Bangladesh. These populations show higher population diversities and higher TB incidence rates compared to the rest of the states. So, in this study we monitored the genetic variants of *NAT2, PXR, GSTs*, and *ABCB1* genes in TB patients to generate baseline data to calculate the risk of developing AT-DILI for better TB management.

## Materials And Method

### Subjects

This study was approved by the ethical committees of Jawaharlal Nehru Institute of Medical Sciences (JNIMS)-Imphal, Manipur (Ref No. Ac/04/IEC/JNIMS/2017), Agartala Govt. Medical College (AGMC)-Agartala, Tripura (Ref. No. F.4(6-9)/AGMC/Academic/IEC Committee/2015/8965 dated 25th April 2018); and Naga Hospital Authority (NHAK)-Kohima, Nagaland (NHAK/HLRC-008/2012 dated 17^th^ May 2017), Manipur University (MU), (Ref No. Ac/IHEC/MU/003/2017) and International Centre for Genetic Engineering and Biotechnology (ICGEB), New Delhi (IEC/IRB No. ICGEB/IEC/2018/06). Subjects reporting with any of the symptoms (>2 weeks of cough, weight loss, night sweat, fever) of TB to the outpatient departments of JNIMS-Imphal, AGMC-Agartala, and NHAK-Kohima representing three Indian northeastern states (Manipur, Tripura and Nagaland) respectively were recruited. After receiving signed informed consent from the study participants, sputum samples were collected for the microscopy, culture and/or GeneXpert tests. And subjects with positive test results of all these tests were selected as active TB(ATB). Whole venous blood samples (~2 ml) were collected in a vial containing EDTA for genomic DNA extraction.

### Genomic DNA Extraction

Whole blood samples were used for genomic DNA (gDNA) extraction using DNA extraction kits (QIAmp DNA Mini Kits, Qiagen) in a QIACube system (Qiagen). Extracted genetic materials were quantified using a spectrophotometer (BioPhotometer, Eppendorf), and integrity was monitored by running agarose gel electrophoresis.

### Genotyping Analysis

A total of six SNPs, three for *NAT2*, and one each for *PXR, ABCB1*, and *GSTT1* genes were selected for the study. Details of the markers used in this study are tabulated in Table 1. Genotyping assays were performed in a qPCR (Rotor-Gene, Qiagen) using TaqMan allelic discrimination probes (Thermo Fisher, USA). The reaction mixture (10 μL) was prepared by adding gDNA (10 ng), TaqMan Universal PCR Master Mix II (5 μL) with uracil N-glycosylase (UNG2X), and Drug Metabolism Genotyping Assay Mix (20X, 0.5 μL) consisting of forward and reverse primer (18 µM) and probes specific for each allele (4 µM). The amplification process involved two steps: a hold at 95 °C for 10 minutes, followed by 50 cycles of amplification including denaturation at 92 °C for 15 seconds and annealing/extension at 60 °C for 90 seconds following the manufacturer’s protocols. The presence or absence of these alleles were analyzed and identified based on the amplification peaks of its probes (VIC/FAM fluorescent dye labeled) specific to the alleles.

**Table 1:**
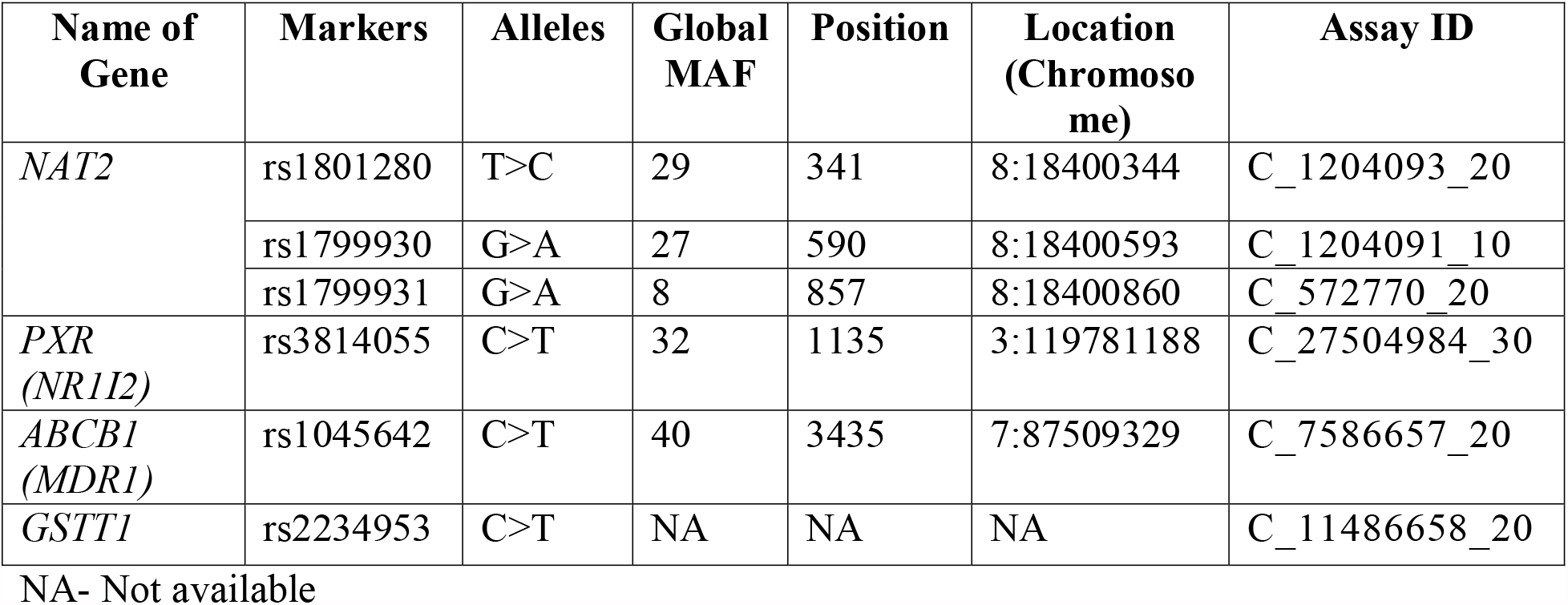
Details of the markers used in this study.

| Name of Gene | Markers | Alleles | Global MAF | Position | Location (Chromosome) | Assay ID |
| --- | --- | --- | --- | --- | --- | --- |
| NAT2 | rs1801280 | T>C | 29 | 341 | 8:18400344 | C_1204093_20 |
|  | rs1799930 | G>A | 27 | 590 | 8:18400593 | C_1204091_10 |
|  | rs1799931 | G>A | 8 | 857 | 8:18400860 | C_572770_20 |
| PXR (NR1I2) | rs3814055 | C>T | 32 | 1135 | 3:119781188 | C_27504984_30 |
| ABCB1 (MDR1) | rs1045642 | C>T | 40 | 3435 | 7:87509329 | C_7586657_20 |
| GSTT1 | rs2234953 | C>T | NA | NA | NA | C_11486658_20 |

NA- Not available

### Statistical analysis

Hardy Weinberg Equilibrium (HWE) analysis was performed for each marker in the population using SPSS software (16.0). Allelic and genotypic frequency were calculated, and a Chi-square test was performed to determine the allelic or genotypic differences among the study populations. Linkage disequilibrium (LD) between the *NAT2* markers was analyzed using the Haploview program (4.1). Genetic distance between populations was estimated using Popgene32 software (1.32). Statistical significance was assumed at *p*≤0.05 at 95% confidence.

## Results

### Patient details

A total of 219 TB patients (male:female, 158/61, mean age (range) in years) from three northeastern states of India (Manipur: 139; Tripura: 60; Nagaland: 20) were included in this study. The mean age of the study population was 46 years.

### Allelic and genotypic distribution of *NAT2*

The *NAT2* gene profile of the study population showed four alleles (NAT2*4, NAT2*5, NAT2*6, and NAT2*7) and ten different genotypes. Genotypic distributions of two of these markers confirmed homogenous distribution following HWE except for NAT2*6 marker (Table 2). Genotypes of three variants of the *NAT2* gene and their corresponding phenotypic profile are tabulated in Table 3. The allelic frequencies of the three variants of *NAT2* in the studied population were found to be 19.9% (95% CI, 1.32-1.48), 24.32% (95% CI, 1.40-1.56), and 18.12% (95% CI, 1.27-1.44) respectively (Table 5). The genotypic frequency of NAT2*4/*6 was highest (~20.1%), and the least was observed for NAT2*5/*7 (~3.2%) as shown in Table 3. The allelic frequencies of *NAT2* markers were significantly (*p*≤0.05) different among the three sub-populations (Table 5). However, no significant difference was observed in the genotypic frequencies among the TB patients from three study sites. Manipur and Nagaland populations have higher frequencies of intermediate acetylator genotypes. NAT2*5/*6, NAT2*5/*7, NAT2*6/*6, and NAT2*6/*7 genotypes were not detected in the Nagaland population (Table 4). The majority of these study populations were found to be intermediate acetylators (~46.1 %) and the rest were either acetylators (~35.6%) or fast acetylators (~17.3 %). Majority (~55%) of Tripura ATB patients showed slow acetylator phenotype, followed by Nagaland (~30%) and Manipur (~27.4%).

**Table 2:** Hardy Weinberg’s Equilibrium analyses of the population in the study. Tripura.

| Marker | Genotype | Chi-square value | P-value |
| --- | --- | --- | --- |
| NAT2*5 | TT | 6.771 | 0.034* |
|  | TC |  |  |
|  | CC |  |  |
| NAT2*6 | GG | 0.198 | 0.906 |
|  | GA |  |  |
|  | AA |  |  |
| NAT2*7 | GG | 24.760 | 0.000* |
|  | GA |  |  |
|  | AA |  |  |
| PXR | CC | 0.237 | 0.888 |
|  | CT |  |  |
|  | TT |  |  |
| ABCB1 | CC | 13.607 | 0.01* |
|  | CT |  |  |
|  | TT |  |  |
| GSTT1 | CC | 56.260 | 0.000* |
|  | NULL |  |  |

**Table 3:**
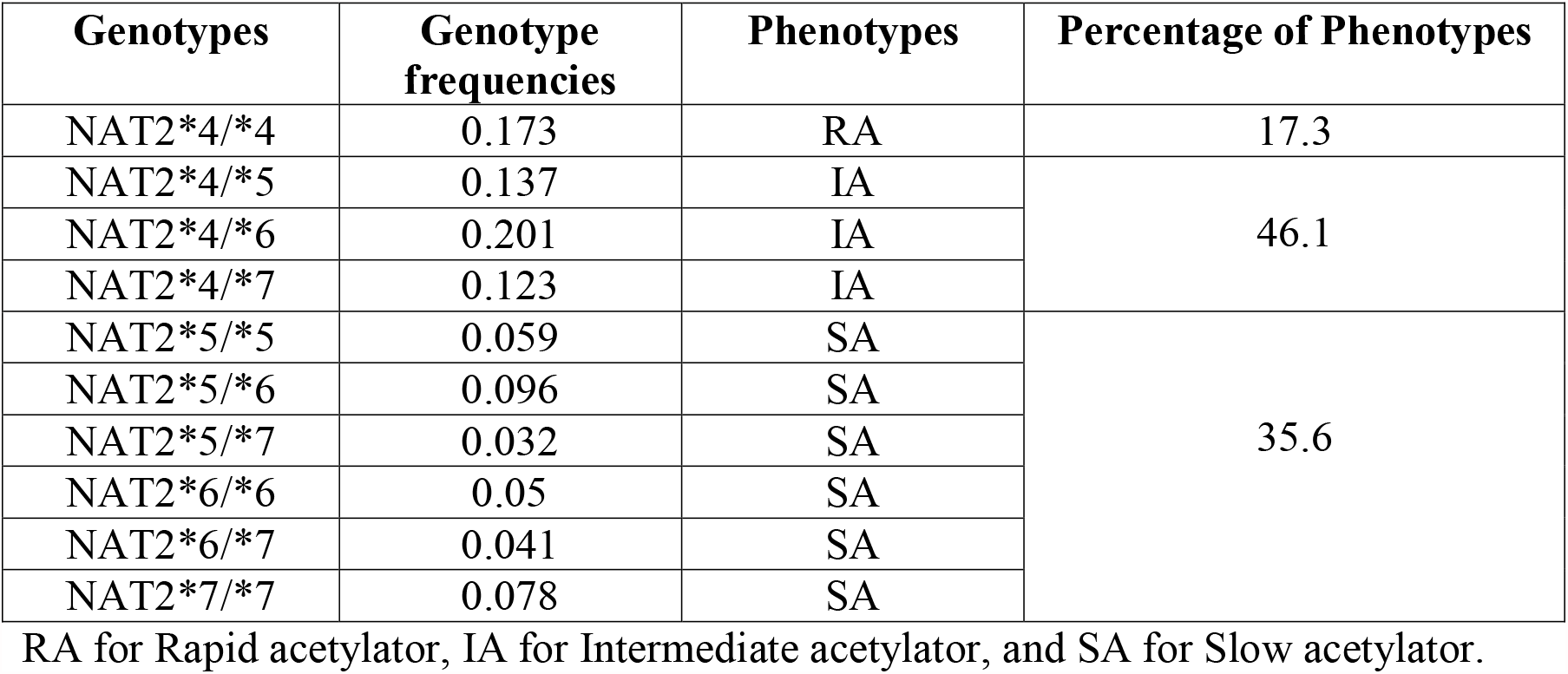
Genotypic frequency of NAT2 gene among the TB patients of Manipur, Nagaland and Tripura.

**Table 4:** Acetylator status of the three study populations.

| Acetylator Status |  |  |  |  |  |  |  |  |  |  |
| --- | --- | --- | --- | --- | --- | --- | --- | --- | --- | --- |
| Genotypes | Fast | Intermediate |  |  |  | Slow |  |  |  |  |
|  | NAT2*<br>4/*4 | NAT2*<br>4/*5 | NAT2*<br>4/*6 | NAT2*<br>4/*7 | NAT2*<br>5/*5 | NAT2*<br>5/*6 | NAT2*<br>5/*7 | NAT2*<br>6/*6 | NAT2*<br>6/*7 | NAT2*<br>7/*7 |
| **Tripura** | 7 (11.7) | 7(11.7) | 12(20) | 01(1.7) | 7(11.7) | 13(21.7) | 02(3.3) | 5(8.3) | 02(3.3) | 4(6.7) |
| Total % of phenotypes | **11.7** | **33.4** |  |  |  | **55** |  |  |  |  |
| **Manipur** | 26(18.7) | 22(15.8) | 29(20.9) | 23(16.5) | 4(2.9) | 8(5.8) | 5(3.6) | 6(4.3) | 6(4.3) | 9(6.5) |
| Total % of phenotypes | **18.7** | **53.2** |  |  |  | **27.4** |  |  |  |  |
| **Nagaland** | 6(30) | 1(5) | 4(20) | 3(15) | 2(10) | 0 | 0 | 0 | 0 | 4(20) |
| Total % of phenotypes | **30** | **40** |  |  |  | **30** |  |  |  |  |

Data shown as: frequency (%)

**Table 5:** Allelic frequencies of NAT2, PXR, ABCB1and GSTT1 among the studied populations.

| Markers<br>(Gene) | Allele | Allelic count in three populations |  |  | Total<br>Population<br>(%) | P-value<br>(95%<br>significa<br>nce) |
| --- | --- | --- | --- | --- | --- | --- |
|  |  | Manipur<br>n=278<br>(freq) | Tripura<br>n=120<br>(freq) | Nagaland<br>n=40<br>(freq) |  |  |
| rs1801280<br>(*NAT2* *5) | T | 234(0.84) | 81(0.675) | 35(0.875) | 80.10 | *0.000 |
|  | C | 44(0.16) | 39(0.325) | 5(0.125) | 19.90 |  |
| rs1799930<br>(*NAT2* *6) | G | 221(0.79) | 76(0.64) | 35(0.875) | 75.68 | *0.000 |
|  | A | 57(0.21) | 44(0.36) | 5(0.125) | 24.32 |  |
| rs1799931<br>(*NAT2* *7) | G | 231(0.83) | 106(0.88) | 28(0.7) | 81.88 | *0.000 |
|  | A | 47(0.17) | 14(0.12) | 12(0.3) | 18.12 |  |
| rs3814055<br>(*PXR*) | C | 231(0.83) | 87(0.725) | 32(0.8) | 80.0 | 0.053 |
|  | T | 47(0.17) | 33(0.275) | 8(0.2) | 20.0 |  |
| rs1045642<br>(*ABCB1*) | C | 114(0.41) | 48(0.4) | 16(0.4) | 41.0 | 0.979 |
|  | T | 164(0.59) | 72(0.6) | 24(0.6) | 59.0 |  |
| rs2234953<br>(*GSTT1*) | C | 194(0.69) | 110(0.92) | 26(0.65) | 75.3 | *0.000 |
|  | Null | 84(0.31) | 10(0.08) | 14(0.35) | 24.7 |  |

### Linkage Disequilibrium (LD) analysis between *NAT2* markers

LD analysis of the three *NAT2* variants, rs1801280 (341 T>C), rs1799930 (590 G>A), and rs1799931 (857 G>A) using Haploview software showed no particular block between the markers. A high LD score (D’=0.82) was observed between rs1801280 and rs1799931 (Fig. 1). The LD strength between rs1801280-rs1799930 and rs1799930-rs1799931 were comparatively lower than rs1801280-rs1799931.

**Fig. 1.**
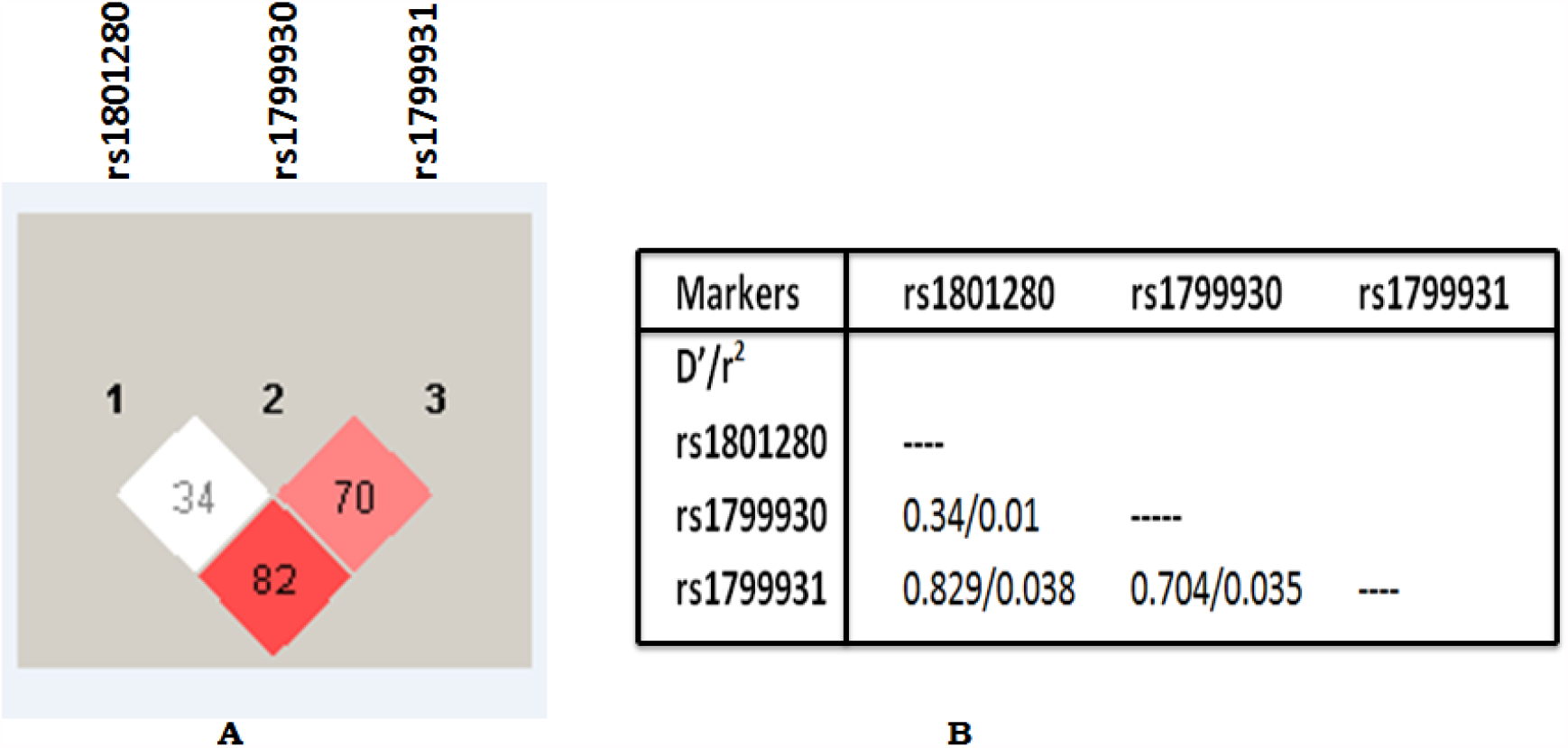
**(A)** Graphical representation of pairwise linkage disequilibrium (LD) of *NAT2* gene markers in TB patients. The numerical values in the boxes represent the normalized linkage disequilibrium coefficient D’ values. The color intensities of the boxes is dependent on the D’/LOD value. The higher the intensity of the red colour, higher is the D’ value. (**B)** LD analysis between marker pairs of *NAT2* gene in TB patients indicating the ratio of D’ to correlation coefficient (D’/r^2^)

### Allelic and genotypic distribution of *PXR*

The *PXR* marker in the study population did not confirm to HWE. The allelic and genotypic frequencies of the *PXR* gene (rs3814055, 25385 C>T) as observed in the study population are tabulated in Table 5 and Table 6. The homozygous C and T genotypes were observed to be ~64.2% and ~4.6%, respectively and ~31.2% was of heterozygous CT genotype. The allelic frequencies of C and T in the study population were found to be ~80% and ~20%, respectively. All three sub-populations showed a higher frequency of homozygous CC genotypes (Tripura: ~51.7%; Manipur: ~69.60%, and Nagaland: ~65%) compared to TT or CT genotypes. No significant differences were observed in the allelic and genotypic frequencies of the *PXR* marker among these three study sites.

**Table 6:**
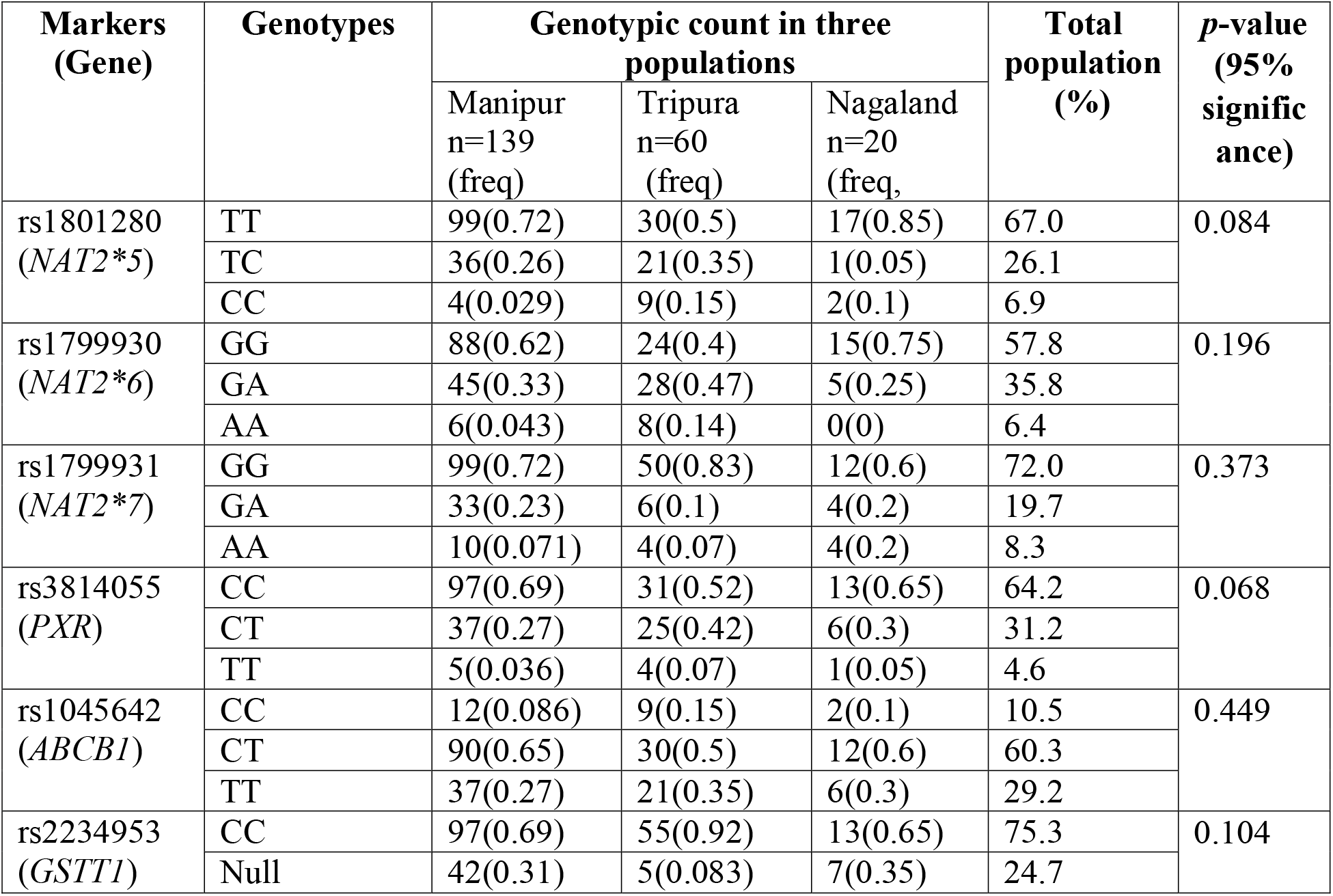
Genotypic frequencies of *NAT2, PXR, ABCB1* and *GSTT1*among TB patients in three populations of North East India.

### Allelic and genotypic distribution of *ABCB1*

The *ABCB1* marker distribution in the study population conforms to HWE (p=0.01, Table 2). The allelic and genotypic frequencies of the *ABCB1* marker are tabulated (Tables 5 and Table 6). No significant differences in the allelic and genotypic frequencies of the marker between study sites were observed in this study. The heterozygous CT genotype was observed to be the highest (~60.3%) compared to TT (~29.2%) or CC (~10.5%) genotypes in the study population. Study populations from all three sites also show ‘T’ to be the more frequent allele (Tripura: ~60%, Manipur: ~59%, Nagaland: ~60%) compared to the ‘C’ allele.

### Allelic and genotypic distribution of *GSTT1*

The *GSTT1* marker in the study population conforms to HWE (p=0.000, Table 2). Homozygous CC genotype was observed to be more prevalent in all three sub-populations (Table 6; Tripura: ~92%, Manipur: 69%, and Nagaland: 65%). ‘C’ allele has higher frequency in the population, and a significant difference in the distribution of the allele was also observed among the populations from three study sites (p<0.00001, Table 5). The null genotype of *GSTT1* marker in the study populations from Tripura, Manipur, and Nagaland were ~8.3%, ~31%, and ~35%, respectively (Table 6).

### Genetic distance of ATB patients between the three study sites

A dendrogram constructed using the frequency of different alleles of *NAT2, PXR, ABCB1*, and *GSTT1* showed that Manipur and Nagaland populations share a closer genetic similarity compared to Tripura population (Fig 2).

**Fig 2:**
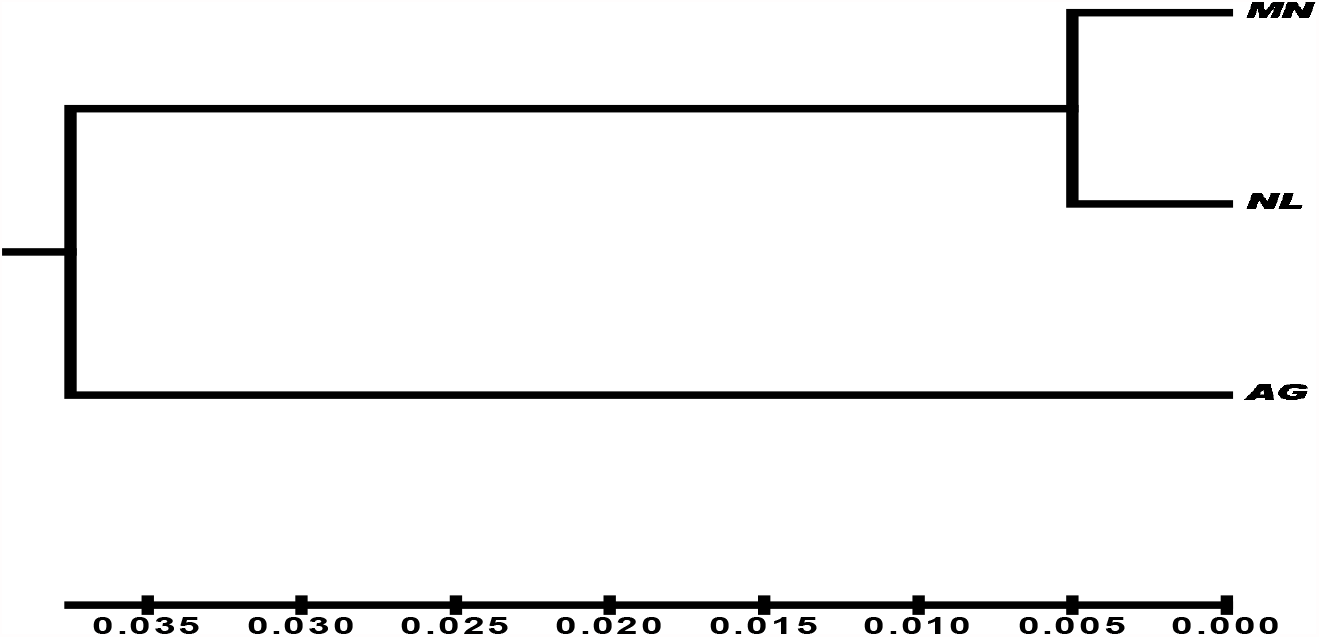
UPGMA dendrogram based Nei’s (1972) unbiased measure of genetic distance among the three study populations representing different north eastern states of India (MN- Manipur; NL-Nagaland; AG- Tripura).

### Combinational analysis of different genotypes

The prevalence of different combinations of the genotypes is reported to be associated with DILI. Both SA genotype of *NAT2* and *GSTT1* null genotype were observed in ~8.2% of the study population. Similarly, ~3.2% of the study population has both SA (*NAT2*) and TT (*PXR*) genotypes. Approximately 10.5% showed both SA (*NAT2*) and TT (*ABCB1)* genotypes. A combination of SA (*NAT2*), TT (*ABCB1)*, and *GSTT1* null genotypes were observed in ~3.7% of the study population. In one of the subjects (~0.5%), a combination of all the four genotypes-SA (*NAT2*), TT (*ABCB1), TT (PXR)*, and *GSTT1* null was detected. Overall, ~26.1% of the study population carried the risk factor genotypes for DILI. Population-wise, ~20.9% of the Manipur population carried the risk factor genotype, ~31.8% in Tripura and ~45% in the Nagaland population.

## Discussion

It is well documented that host genetics plays a crucial role in metabolizing xenobiotic compounds including drugs and it significantly influence therapeutic outcomes. In case of infectious diseases like TB in which patients undertake medication for a longer duration (6-18 months) may develop severe side effects like hepatotoxicity, nephrotoxicity due to incomplete metabolism and clearance of drugs or their intermediate products. In this study, we aimed to generate baseline pharmacogenomics data of understudied TB patient populations from three different north-eastern states of India sharing international boundaries with Myanmar and Bangladesh.

It is well known that NAT2 polymorphism impacts drug metabolism and based on their patterns, the subjects could be grouped as slow-, intermediate- or fast-acetylators. The majority of this study population were found to have *NAT2*4/*6* genotype, representing intermediate acetylator types. Similar observations were earlier reported from one of the neighboring state i.e. Assam [39]. NAT2*6 allele, which has been associated with a risk for DILI, was found to be the most common in this study population and corroborates earlier findings [40,14]. However, in the Tripura population, we observed the majority of the study subjects present with the slow-acetylator phenotypes. This genomics data support our earlier metabolomics related findings in which the abundance of urine anti-TB drugs and their intermediate levels were monitored using mass spectrometry and demonstrated their slow acetylation status [41]. This could be partly explained by the Indo-Aryan origin of the Tripura population whereas the Manipur and Nagaland study population sharing a common East Asian origin.

The allelic frequencies of the three variants of *NAT2* in the studied population were ~19.9%, ~24.32%, and ~18.12% respectively, which is different from earlier reports [40, 41]. This could be explained by the difference in allelic frequency of the *NAT2* variants with ethnicity (Table 7).

**Table 7:** *NAT2* allele frequencies among Indian population and other ethnic groups.

| Populations | Frequency (%) |  |  |  |  |
| --- | --- | --- | --- | --- | --- |
|  | n | 341 T>C | 590 G>A | 857 G>A | references |
| Indian | 250 | 15.6 | 42.8 | 8.4 | [39] |
| Indians | 61 | 33.0 | 38 | 3.0 | [40] |
| Japanese | 79 | 1.9 | 23 | 1.1 | [40] |
| Caucasians | 3531 | 46.0 | 28.5 | 2.9 | [42] |
| Southern Korean | 288 | 1.0 | 22.4 | 13.2 | [43] |
| Americans | 387 | 4.37 | 26.6 | 1.9 | [43] |
| Southern Brazil | 254 | 28.9 | 10.4 | 2.1 | [44] |
| South Africa | 97 | 36.1 | 17.0 | 6.7 | [43] |
| Spanish | 258 | 47 | 25.0 | 0.6 | [43] |
| Egyptian | 199 | 49.7 | 26 | 2.8 | [45] |
| Tunisians | 100 | 31.5 | 17.5 | 15 | [46] |
| **North East Indian (Current study)** | **219** | **19.9** | **24.32** | **18.12** | **Current study** |

In the study population, we observed higher ancestral C alleles of the PXR gene and homozygous CC genotypes than the variant T alleles. In the case of the *ABCB1 (MDR-1)* gene, T alleles are more prevalent than the C allele in the studied population. A higher prevalence of the heterozygous CT genotypes was observed than the homozygous TT genotype corroborating earlier reports (Table 8). In an earlier study on asian population (**n=298**, from Singapore), Balram et al., reported a C and T allele frequencies of ~38% and ~62% and similar observations were observed in our study population (C: 41% -, T: 59%) [47, 52, 54]. However, the earlier reported study population had a higher prevalence of the homozygous TT genotype than the heterozygous CT genotype and may be due to the differences in ethnicity.

**Table 8:** Genotype and allele frequencies of *ABCB1* (*MDR-1)* gene C3435T polymorphism in various ethnic groups.

| Populations (n) | n | CC | CT | TT | C | T | references |
| --- | --- | --- | --- | --- | --- | --- | --- |
| Indians | 93 | 18.0 | 39.0 | 43.0 | 0.380 | 0.620 | [47] |
| Jordanians | 100 | 17.0 | 50.0 | 33.0 | 0.420 | 0.580 | [48] |
| Iranians | 200 | 20.0 | 52.5 | 27.5 | 0.463 | 0.537 | [49] |
| Turkish | 150 | 20.0 | 53.0 | 27.0 | 0.470 | 0.530 | [50] |
| Spanish | 408 | 26.0 | 52.0 | 22.0 | 0.520 | 0.480 | [51] |
| British | 190 | 24.0 | 48.0 | 28.0 | 0.480 | 0.520 | [52] |
| Chinese | 132 | 32.0 | 42.0 | 26.0 | 0.530 | 0.470 | [52] |
| German | 188 | 28.0 | 48.0 | 24.0 | 0.520 | 0.480 | [36] |
| Japanese | 154 | 35.7 | 47.4 | 16.9 | 0.594 | 0.406 | [53] |
| Ghanaians | 206 | 66.0 | 34.0 | 0 | 0.830 | 0.170 | [52] |
| French | 81 | 36.0 | 42.0 | 22.0 | 0.570 | 0.430 | [54] |
| **Current study** | **219** | **10.5** | **60.3** | **29.2** | **0.41** | **0.59** | **Current study** |

A higher prevalence of homozygous CC than the null genotypes for *GSTT1* was observed in this study population. Homozygous null mutation at this locus in Manipur (~31%) and Nagaland (~35%) populations were higher than Tripura (~8.3%) population. Overall, ~75% of the population in the study has a homozygous CC genotype, exhibiting limited loss of Glutathione S-transferase activity in this study population. From the genetic distance analysis, the population from Manipur and Nagaland were found to be closer to each other compared to the Tripura population (Fig. 2).

Analysis of different combinations of the genotypes associated with DILI such as slow acetylator (SA) genotype of *NAT2, GSTT1* null genotype, homozygous TT genotype of *PXR* and homozygous TT genotype of *ABCB1* (Table 9) showed that ~26.1 % of the study population has two or more combination of these risk genotypes. Earlier reports demonstrated combinations of SA genotype of *NAT2* and null genotypes of GSTs were significantly associated with AT-DILI. A single study subject (~0.5%) from Manipur, exhibited all the four genotypes associated with DILI. In the Manipur population, ~20.9% showed different combinations of risk genotypes. Nagaland (~45%) and Tripura (~31.9%) populations showed different combinations of risk genotypes. With this relatively small sample size, the baseline data was generated from three study sites, on which limited or no prior data on pharmacogenomics was available. Further focused expansion of this study will bring additional clarity on these understudied populations. Overall, the prevalence of the risk factor genotypes of the studied genes is ~ 26.1%, which indicates a low risk to develop DILI during TB treatment in these patients.

**Table 9:**
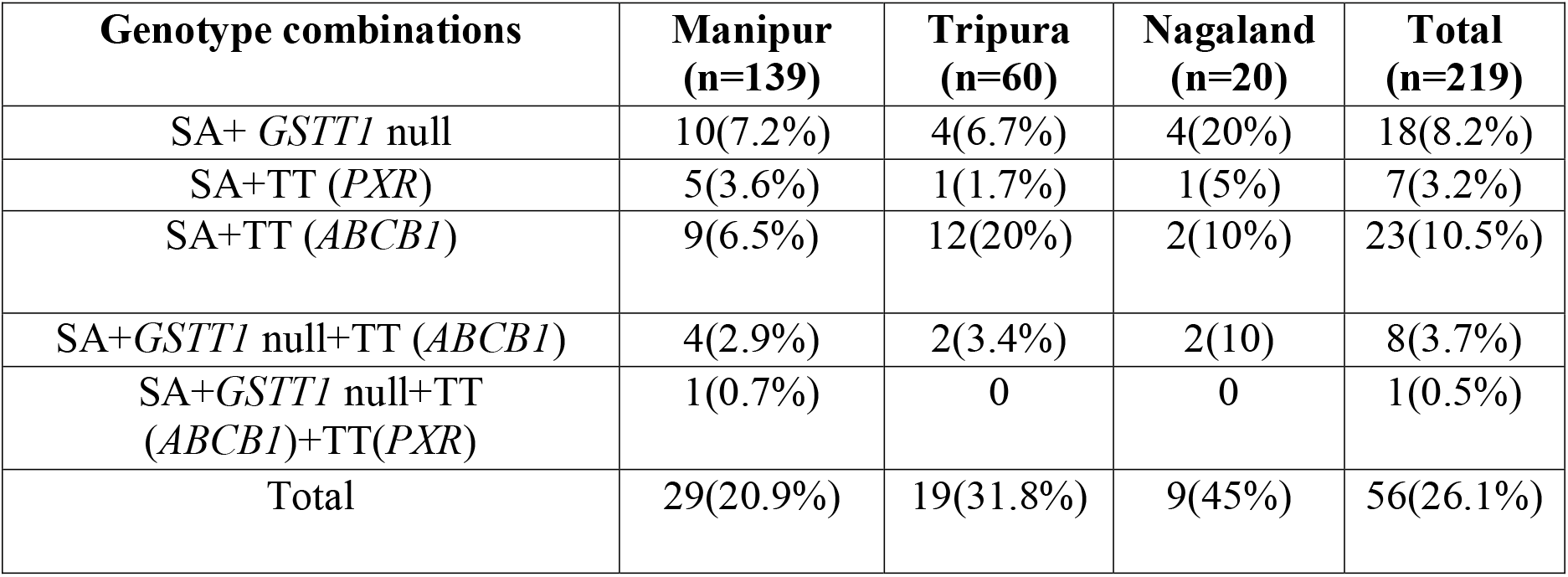
Analysis of different combinations of risk genotypes of *NAT2, PXR, GSTT1*, and *ABCB1* genes associated with TB-DILI in the three populations of the study.

In conclusion, this pilot genetic polymorphism study including *NAT2, PXR, GSTs*, and *ABCB1* genes in TB patients from the three north-eastern states of India showed that majority of these patients were carriers of ancestral allele and genotypes with low risk to DILI development during the treatment. This baseline data could be expanded further with additional marker profiles and population size to develop appropriate policy interventions for these under-studied areas.

## Data Availability

The data related to the manuscript are available with the authors.

## Acknowledgements

We are thankful to all the study participants and clinical staff of all partnering sites involved during patient recruitment for their support.

## Ethics approval

This study was approved by the ethical committees of Jawaharlal Nehru Institute of Medical Sciences (JNIMS)-Imphal, Manipur (Ref No. Ac/04/IEC/JNIMS/2017), Agartala Govt. Medical College (AGMC)-Agartala, Tripura (Ref. No. F.4(6-9)/AGMC/Academic/IEC Committee/2015/8965 dated 25th April 2018); and Naga Hospital Authority (NHAK)-Kohima, Nagaland (NHAK/HLRC-008/2012 dated 17^th^ May 2017), Manipur University (MU), (Ref No. Ac/IHEC/MU/003/2017) and International Centre for Genetic Engineering and Biotechnology (ICGEB), New Delhi (IEC/IRB No. ICGEB/IEC/2018/06).

## Consent to participate

Signed informed consent forms to participate in this study were received from the participants before enrolling in this study.

## Consent for publication

All contributing authors have gone through the manuscript and provided consent for submitting this manuscript for publication.

## References

1. https://www.who.int/publications/i/item/9789240013131 (accessed on 7th June 2021)

2. https://tbcindia.gov.in/WriteReadData/l892s/India%20TB%20Report%202020.pdf (accessed on 7th June 2021)

3. Martinjak-Dvorsek I, Gorjup V, Horvat M, Noc M. Acute isoniazid neurotoxicity during preventive therapy. Crit Care Med. 2000 Feb;28(2):567–8.

4. Watkins RC, Hambrick EL, Benjamin G, Chavda SN. Isoniazid toxicity presenting as seizures and metabolic acidosis. J Natl Med Assoc. 1990 Jan;82(1):57, 62, 64.

5. Higuchi N, Tahara N, Yanagihara K, Fukushima K, Suyama N, Inoue Y, Miyazaki Y, Kobayashi T, Yoshiura K, Niikawa N, Wen CY, Isomoto H, Shikuwa S, Omagari K, Mizuta Y, Kohno S, Tsukamoto K. NAT2 6A, a haplotype of the N-acetyltransferase 2 gene, is an important biomarker for risk of anti-tuberculosis drug-induced hepatotoxicity in Japanese patients with tuberculosis. World J Gastroenterol. 2007 Dec 7;13(45):6003–8.

6. Shu CC, Lee CH, Lee MC, Wang JY, Yu CJ, Lee LN. Hepatotoxicity due to first-line anti-tuberculosis drugs: a five-year experience in a Taiwan medical centre. Int J Tuberc Lung Dis. 2013 Jul;17(7):934–9.

7. Pande JN, Singh SP, Khilnani GC, Khilnani S, Tandon RK. Risk factors for hepatotoxicity from antituberculosis drugs: a case-control study. Thorax. 1996 Feb;51(2):132–6.

8. Wu JC, Lee SD, Yeh PF, Chan CY, Wang YJ, Huang YS, Tsai YT, Lee PY, Ting LP, Lo KJ. Isoniazid-rifampin-induced hepatitis in hepatitis B carriers. Gastroenterology. 1990 Feb;98(2):502–4.

9. Durand F, Bernuau J, Pessayre D, Samuel D, Belaiche J, Degott C, Bismuth H, Belghiti J, Erlinger S, Rueff B, et al. Deleterious influence of pyrazinamide on the outcome of patients with fulminant or subfulminant liver failure during antituberculous treatment including isoniazid. Hepatology. 1995 Apr;21(4):929–32.

10. Singh J, Garg PK, Thakur VS, Tandon RK. Anti tubercular treatment induced hepatotoxicity: does acetylator status matter? Indian J Physiol Pharmacol. 1995 Jan;39(1):43–6.

11. Wong WM, Wu PC, Yuen MF, Cheng CC, Yew WW, Wong PC, Tam CM, Leung CC, Lai CL. Antituberculosis drug-related liver dysfunction in chronic hepatitis B infection. Hepatology. 2000 Jan;31(1):201–6.

12. Ungo JR, Jones D, Ashkin D, Hollender ES, Bernstein D, Albanese AP, Pitchenik AE. Antituberculosis drug-induced hepatotoxicity. The role of hepatitis C virus and the human immunodeficiency virus. Am J Respir Crit Care Med. 1998 Jun;157(6 Pt 1):1871–6.

13. Shakya R, Rao BS, Shrestha B. Incidence of hepatotoxicity due to antitubercular medicines and assessment of risk factors. Ann Pharmacother. 2004 Jun;38(6):1074–9.

14. Suvichapanich S, Wattanapokayakit S, Mushiroda T, Yanai H, Chuchottawon C, Kantima T, Nedsuwan S, Suwankesawong W, Sonsupap C, Pannarunothai R, Tumpattanakul S, Bamrungram W, Chaiwong A, Mahasirimongkol S, Mameechai S, Panthong W, Klungtes N, Munsoo A, Chauychana U, Maneerat M, Fukunaga K, Omae Y, Tokunaga K. Genomewide Association Study Confirming the Association of NAT2 with Susceptibility to Antituberculosis Drug-Induced Liver Injury in Thai Patients. Antimicrob Agents Chemother. 2019 Jul 25;63(8):e02692–18.

15. Bao Y, Ma X, Rasmussen TP, Zhong XB. Genetic Variations Associated with Anti-Tuberculosis Drug-Induced Liver Injury. Curr Pharmacol Rep. 2018 Jun;4(3):171–181.

16. Ohno M, Yamaguchi I, Yamamoto I, Fukuda T, Yokota S, Maekura R, Ito M, Yamamoto Y, Ogura T, Maeda K, Komuta K, Igarashi T, Azuma J. Slow N-acetyltransferase 2 genotype affects the incidence of isoniazid and rifampicin-induced hepatotoxicity. Int J Tuberc Lung Dis. 2000 Mar;4(3):256–61.

17. Huang YS, Chern HD, Su WJ, Wu JC, Chang SC, Chiang CH, Chang FY, Lee SD. Cytochrome P450 2E1 genotype and the susceptibility to antituberculosis drug-induced hepatitis. Hepatology. 2003 Apr;37(4):924–30.

18. Sharma SK, Balamurugan A, Saha PK, Pandey RM, Mehra NK. Evaluation of clinical and immunogenetic risk factors for the development of hepatotoxicity during antituberculosis treatment. Am J Respir Crit Care Med. 2002 Oct 1;166(7):916–9.

19. Shimizu Y, Dobashi K, Mita Y, Endou K, Moriya S, Osano K, Koike Y, Higuchi S, Yabe S, Utsugi M, Ishizuka T, Hisada T, Nakazawa T, Mori M. DNA microarray genotyping of N-acetyltransferase 2 polymorphism using carbodiimide as the linker for assessment of isoniazid hepatotoxicity. Tuberculosis (Edinb). 2006 Sep;86(5):374–81.

20. Parkin DP, Vandenplas S, Botha FJ, Vandenplas ML, Seifart HI, van Helden PD, van der Walt BJ, Donald PR, van Jaarsveld PP. Trimodality of isoniazid elimination: phenotype and genotype in patients with tuberculosis. Am J Respir Crit Care Med. 1997 May;155(5):1717–22.

21. Mitchell JR, Thorgeirsson UP, Black M, Timbrell JA, Snodgrass WR, Potter WZ, Jollow HR, Keiser HR. Increased incidence of isoniazid hepatitis in rapid acetylators: possible relation to hydranize metabolites. Clin Pharmacol Ther. 1975 Jul;18(1):70–9.

22. Cho HJ, Koh WJ, Ryu YJ, Ki CS, Nam MH, Kim JW, Lee SY. Genetic polymorphisms of NAT2 and CYP2E1 associated with antituberculosis drug-induced hepatotoxicity in Korean patients with pulmonary tuberculosis. Tuberculosis (Edinb). 2007 Nov;87(6):551–6.

23. Zang Y, Doll MA, Zhao S, States JC, Hein DW. Functional characterization of single-nucleotide polymorphisms and haplotypes of human N-acetyltransferase 2. Carcinogenesis. 2007 Aug;28(8):1665–71.

24. Cai Y, Yi J, Zhou C, Shen X. Pharmacogenetic study of drug-metabolising enzyme polymorphisms on the risk of anti-tuberculosis drug-induced liver injury: a meta-analysis. PLoS One. 2012;7(10):e47769.

25. Wang PY, Xie SY, Hao Q, Zhang C, Jiang BF. NAT2 polymorphisms and susceptibility to anti-tuberculosis drug-induced liver injury: a meta-analysis. Int J Tuberc Lung Dis. 2012 May;16(5):589–95.

26. Ng CS, Hasnat A, Al Maruf A, Ahmed MU, Pirmohamed M, Day CP, Aithal GP, Daly AK. N-acetyltransferase 2 (NAT2) genotype as a risk factor for development of drug-induced liver injury relating to antituberculosis drug treatment in a mixed-ethnicity patient group. Eur J Clin Pharmacol. 2014 Sep;70(9):1079–86.

27. Singla N, Gupta D, Birbian N, Singh J. Association of NAT2, GST and CYP2E1 polymorphisms and anti-tuberculosis drug-induced hepatotoxicity. Tuberculosis (Edinb). 2014 May;94(3):293–8.

28. Tolson AH, Wang H. Regulation of drug-metabolizing enzymes by xenobiotic receptors: PXR and CAR. Adv Drug Deliv Rev. 2010 Oct 30;62(13):1238–49.

29. Moore LB, Goodwin B, Jones SA, Wisely GB, Serabjit-Singh CJ, Willson TM, Collins JL, Kliewer SA. St. John’s wort induces hepatic drug metabolism through activation of the pregnane X receptor. Proc Natl Acad Sci U S A. 2000 Jun 20;97(13):7500–2.

30. Jancova P, Anzenbacher P, Anzenbacherova E. Phase II drug metabolizing enzymes. Biomed Pap Med Fac Univ Palacky Olomouc Czech Repub. 2010 Jun;154(2):103–16.

31. Zazuli Z, Barliana MI, Mulyani UA, Perwitasari DA, Ng H, Abdulah R. Polymorphism of PXR gene associated with the increased risk of drug-induced liver injury in Indonesian pulmonary tuberculosis patients. J Clin Pharm Ther. 2015 Dec;40(6):680–4.

32. Leiro V, Fernández-Villar A, Valverde D, Constenla L, Vázquez R, Piñeiro L, González-Quintela Influence of glutathione S-transferase M1 and T1 homozygous null mutations on the risk of antituberculosis drug-induced hepatotoxicity in a Caucasian population. Liver Int. 2008 Jul;28(6):835–9.

33. Roy B, Chowdhury A, Kundu S, Santra A, Dey B, Chakraborty M, Majumder PP. Increased risk of antituberculosis drug-induced hepatotoxicity in individuals with glutathione S-transferase M1 ‘null’ mutation. J Gastroenterol Hepatol. 2001 Sep;16(9):1033–7.

34. Naumovska Z, Nestorovska AK, Sterjev Z, Filipce A, Dimovski A, Suturkova L. Genotype variability and haplotype profile of ABCB1 (MDR1) gene polymorphisms in Macedonian population. Pril (Makedon Akad Nauk Umet Odd Med Nauki). 2014;35(3):121–33.

35. Komar AA. Silent SNPs: impact on gene function and phenotype. Pharmacogenomics. 2007 Aug;8(8):1075–80.

36. Hoffmeyer S, Burk O, von Richter O, Arnold HP, Brockmöller J, Johne A, Cascorbi I, Gerloff T, Roots I, Eichelbaum M, Brinkmann U. Functional polymorphisms of the human multidrug-resistance gene: multiple sequence variations and correlation of one allele with P-glycoprotein expression and activity in vivo. Proc Natl Acad Sci U S A. 2000 Mar 28;97(7):3473–8.

37. Lu PH, Wei MX, Yang J, Liu X, Tao GQ, Shen W, Chen MB. Association between two polymorphisms of ABCB1 and breast cancer risk in the current studies: a meta-analysis. Breast Cancer Res Treat. 2011 Jan;125(2):537–43.

38. Yimer G, Ueda N, Habtewold A, Amogne W, Suda A, Riedel KD, Burhenne J, Aderaye G, Lindquist L, Makonnen E, Aklillu E. Pharmacogenetic & pharmacokinetic biomarker for efavirenz based ARV and rifampicin based anti-TB drug induced liver injury in TB-HIV infected patients. PLoS One. 2011;6(12):e27810.

39. Khan N, Pande V, Das A. NAT2 sequence polymorphisms and acetylation profiles in Indians. Pharmacogenomics. 2013 Feb;14(3):289–303.

40. Lin HJ, Han CY, Lin BK, Hardy S. Ethnic distribution of slow acetylator mutations in the polymorphic N-acetyltransferase (NAT2) gene. Pharmacogenetics. 1994 Jun;4(3):125–34.

41. Das MK, Arya R, Debnath S, Debnath R, Lodh A, Bishwal SC, Das A, Nanda RK. Global Urine Metabolomics in Patients Treated with First-Line Tuberculosis Drugs and Identification of a Novel Metabolite of Ethambutol. Antimicrob Agents Chemother. 2016 Mar 25;60(4):2257–64.

42. Garte S, Gaspari L, Alexandrie AK, Ambrosone C, Autrup H, Autrup JL, Baranova H, Bathum L, Benhamou S, Boffetta P, Bouchardy C, Breskvar K, Brockmoller J, Cascorbi I, Clapper ML, Coutelle C, Daly A, Dell’Omo M, Dolzan V, Dresler CM, Fryer A, Haugen A, Hein DW, Hildesheim A, Hirvonen A, Hsieh LL, Ingelman-Sundberg M, Kalina I, Kang D, Kihara M, Kiyohara C, Kremers P, Lazarus P, Le Marchand L, Lechner MC, van Lieshout EM, London S, Manni JJ, Maugard CM, Morita S, Nazar-Stewart V, Noda K, Oda Y, Parl FF, Pastorelli R, Persson I, Peters WH, Rannug A, Rebbeck T, Risch A, Roelandt L, Romkes M, Ryberg D, Salagovic J, Schoket B, Seidegard J, Shields PG, Sim E, Sinnet D, Strange RC, Stücker I, Sugimura H, To-Figueras J, Vineis P, Yu MC, Taioli E. Metabolic gene polymorphism frequencies in control populations. Cancer Epidemiol Biomarkers Prev. 2001 Dec;10(12):1239–48.

43. Sabbagh A, Langaney A, Darlu P, Gérard N, Krishnamoorthy R, Poloni ES. Worldwide distribution of NAT2 diversity: implications for NAT2 evolutionary history. BMC Genet. 2008 Feb 27;9:21.

44. Teixeira RL, Morato RG, Cabello PH, Muniz LM, Moreira Ada S, Kritski AL, Mello FC, Suffys PN, Miranda AB, Santos AR. Genetic polymorphisms of NAT2, CYP2E1 and GST enzymes and the occurrence of antituberculosis drug-induced hepatitis in Brazilian TB patients. Mem Inst Oswaldo Cruz. 2011 Sep;106(6):716–24.

45. Hamdy SI, Hiratsuka M, Narahara K, Endo N, El-Enany M, Moursi N, Ahmed MS, Mizugaki M. Genotype and allele frequencies of TPMT, NAT2, GST, SULT1A1 and MDR-1 in the Egyptian population. Br J Clin Pharmacol. 2003 Jun;55(6):560–9.

46. Bendjemana K, Abdennebi M, Gara S, Jmal A, Ghanem A, Touati S, Boussen H, Ladgham A, Guemira F. Genetic polymorphism of gluthation-S transferases and N-acetyl transferases 2 and nasopharyngeal carcinoma: the Tunisia experience. Bull Cancer. 2006 Mar 1;93(3):297–302. French.

47. Balram C, Sharma A, Sivathasan C, Lee EJ. Frequency of C3435T single nucleotide MDR1 genetic polymorphism in an Asian population: phenotypic-genotypic correlates. Br J Clin Pharmacol. 2003 Jul;56(1):78–83.

48. Khabour OF, Alzoubi KH, Al-Azzam SI, Mhaidat NM. Frequency of MDR1 single nucleotide polymorphisms in a Jordanian population, including a novel variant. Genet Mol Res. 2013 Mar 13;12(1):801–8.

49. Azarpira N, Aghdaie M H. Frequency of C3435 MDR1 and A6896G CYP3A5 Single Nucleotide Polymorphism in an Iranian Population and Comparison with Other Ethnic Groups. Medical Journal of the Islamic Republic of Iran (2006);20(3);131–136.

50. Turgut S, Turgut G, Atalay EO. Genotype and allele frequency of human multidrug resistance (MDR1) gene C3435T polymorphism in Denizli province of Turkey. Mol Biol Rep. 2006 Dec;33(4):295–300.

51. Bernal ML, Sinues B, Fanlo A, Mayayo E. Frequency distribution of C3435T mutation in exon 26 of the MDR1 gene in a Spanish population. Ther Drug Monit. 2003 Feb;25(1):107–11.

52. Ameyaw MM, Regateiro F, Li T, Liu X, Tariq M, Mobarek A, Thornton N, Folayan GO, Githang’a J, Indalo A, Ofori-Adjei D, Price-Evans DA, McLeod HL. MDR1 pharmacogenetics: frequency of the C3435T mutation in exon 26 is significantly influenced by ethnicity. Pharmacogenetics. 2001 Apr;11(3):217–21.

53. Igumnova V, Capligina V, Krams A, Cirule A, Elferts D, Pole I, Jansone I, Bandere D, Ranka R. Genotype and allele frequencies of isoniazid-metabolizing enzymes NAT2 and GSTM1 in Latvian tuberculosis patients. J Infect Chemother. 2016 Jul;22(7):472–7.

54. Teixeira RL, Morato RG, Cabello PH, Muniz LM, Moreira Ada S, Kritski AL, Mello FC, Suffys PN, Miranda AB, Santos AR. Genetic polymorphisms of NAT2, CYP2E1 and GST enzymes and the occurrence of antituberculosis drug-induced hepatitis in Brazilian TB patients. Mem Inst Oswaldo Cruz. 2011 Sep;106(6):716–24.

